# Clinical, immunological and virological characterization of COVID-19 patients that test re-positive for SARS-CoV-2 by RT-PCR

**DOI:** 10.1101/2020.06.15.20131748

**Authors:** Jing Lu, Jinju Peng, Qianling Xiong, Zhe Liu, Huifang Lin, Xiaohua Tan, Min Kang, Runyu Yuan, Lilian Zeng, Pingping Zhou, Chumin Liang, Lina Yi, Louis du Plessis, Tie Song, Wenjun Ma, Jiufeng Sun, Oliver G Pybus, Changwen Ke

**Affiliations:** Guangdong Provincial Institution of Public Health, Guangdong Provincial Center for Disease Control and Prevention, Guangzhou, China; Guangdong Provincial Center for Disease Control and Prevention, Guangzhou, China; School of Public Health, Southern Medical University, Guangzhou, China; Department of Zoology, University of Oxford, Oxford, UK

**Keywords:** Re-positive, SARS-CoV-2, COVID-19, Neutralizing antibody, Virology

## Abstract

**Background:** COVID-19 pandemic is underway. Some COVID-19 cases re-tested positive for SARS-CoV-2 RNA after discharge raising the public concern on their infectivity. Characterization of re-positive cases are urgently needed for designing intervention strategies.

**Methods:** Clinical data were obtained through Guangdong COVID-19 surveillance network. Neutralization antibody titre was determined using a microneutralization assay. Potential infectivity of clinical samples was evaluated after the cell inoculation. SARS-CoV-2 RNA was detected using three different RT-PCR kits and multiplex PCR with nanopore sequencing.

**Results:** Among 619 discharged COVID-19 cases, 87 were re-tested as SARS-CoV-2 positive in circumstance of social isolation. All re-positive cases had mild or moderate symptoms in initial diagnosis and a younger age distribution (mean, 30.4). Re-positive cases (n=59) exhibited similar neutralization antibodies (NAbs) titre distributions to other COVID-19 cases (n=150) parallel-tested in this study. No infective viral strain could be obtained by culture and none full-length viral genomes could be sequenced for all re-positive cases.

**Conclusions:** Re-positive SARS-CoV-2 was not caused by the secondary infection and was identified in around 14% of discharged cases. A robust Nabs response and a potential virus genome degradation were detected from nearly all re-positive cases suggesting a lower transmission risk, especially through a respiratory route.

## Introduction

Coronavirus disease 2019 (COVID-19) is caused by a novel coronavirus, severe acute respiratory syndrome coronavirus 2 (SARS-CoV-2) (1), which shares a 96% genetic similarity with the most closely related bat origin SARS-like virus (RaTG13) (2). As a newly emergent virus, the clinical features of SARS-CoV-2 infections were largely unknown at the beginning of the outbreak but are becoming gradually clearer as a result of global clinical studies (3, 4). The design of risk assessments and successful interventions for COVID-19 are dependent on how well we understand the course of SARS-CoV-2 infections.

The COVID-19 pandemic is underway (5). Social measures for monitoring, controlling and treating COVID-19 are varied among countries. It has been suggested that the detection of antibodies to SARS-CoV-2 could serve as the basis for an “immunity passport”, however it is currently unclear whether recovered COVID-19 cases have neutralizing antibodies that protecting them from a second infection. There have been reports that some recovered COVID-19 cases have re-tested positive for SARS-CoV-2 RNA a few days after discharge (6, 7). Since a RT-PCR test that targets a short fragment of the virus genome cannot indicate if an individual is infectious or not, we define in this study these observations as “re-positive” cases, not relapse or repeat infection cases.

Re-positive detection of SARS-CoV-2 RNA raises questions about the transmission risk of the disease, including, (i) the percentage of re-positive in COVID-19 discharged cases and its association with clinical characters, (ii) the immune status of re-positive cases, and (iii) infectivity of re-positive cases.

Guangdong Province reported the highest number of COVID-19 cases in China except Hubei. Guangdong launched an enhanced surveillance network and a series of intervention measures in response to the outbreak soon after the first COVID-19 case was reported in December 2019 (8). Since 23 January, all discharged COVID-19 cases were isolated in designated hotels under medical observation for another 14 days. In this study, we screened 619 recovered COVID-19 cases in Guangdong discharged between 23 January and 19 February. One hundred thirty-seven swabs and 59 serum samples from 70 re-positive cases were collected in order to reveal the immunological and virologic characteristics of the SARS-CoV-2 re-positive cases.

## Methods

### Discharge criteria and after discharge measures for COVID-19 cases in Guangdong

Guangdong follows the Diagnosis and Treatment Scheme of SARS-CoV-2 released by the National Health Commission of China with a little modification. The Discharge criteria a for COVID-19 cases in Guangdong include: 1) Body temperature is back to normal for more than three days; 2) Respiratory symptoms improve obviously; 3) Pulmonary imaging shows obvious absorption of inflammation, and 4) Nuclei acid tests negative twice consecutively on both respiratory tract samples such as sputum and nasopharyngeal swabs and digestive tract samples such as stool and anal swabs (sampling interval being at least 24 hours). The interventions measures for discharged COVID-19 cases include: 1) all discharged COVID-19 cases are isolated in designated hotels for another 14 days; 2) during the isolation, discharged cases live in well-ventilated single room, separate dinning, practice hand hygiene and minimize close contact with others; 3) health status are monitored during the isolation and SARS-CoV-2 RT-PCR tests are performed on 7^th^ and 14^th^ or more frequently after discharge; 4) cases could back home only when nuclei acid tests are negative on both respiratory tract samples and digestive tract samples during the isolation. The clinical outcome was categorized as mild, moderate, severe, and critical as previously described (8).

### Case definition and specimen collection

The term “re-positive case” in this study refers to the discharged cases who retested as SARS-CoV-2 positive using real-time Reverse-Transcription Polymerase Chain Reaction (RT-PCR; see below). In Guangdong, all discharged COVID-19 cases were continuously isolated in designated hotels and samples including nasopharyngeal swabs, throat swabs and anal swabs, were collected for RT-PCR diagnosis at 7 days and 14 days after discharge, or more frequently. The demographic, clinical and laboratory information of all confirmed COVID-19 cases were retrieved from Guangdong Provincial COVID-19 surveillance network.

### Viral isolation and RT-PCR

Vero E6 cells were inoculated with 100 µl processed patient sample. Cytopathic effect (CPE) were observed daily. If there was no CPE observed, cell lysis was collected by centrifugation after three repeated freeze-thaw and 100 µl supernatant were used for the second round of passage. For RT-PCR diagnosis, total RNA was extracted from clinical specimens using the QIAamp Viral RNA mini kit (QIAGEN, Germany) according to the manufacturer’s instructions. In this study, 3 RT-PCR kits were used to conduct nucleic acid testing, in an attempt to avoid the occurrence of false negatives. Kit A (DAAN GENE, Guangzhou, China) and Kit B (BioGerm, Shanghai, China) have primers and probes targeting the open reading frame (ORF1ab) and nucleocapsid protein (N), respectively. Kit C (Liferiver, Shanghai, China) is designed to detect RNA-dependent RNA polymerase (RdRp), envelope protein (E) and N.

### Microneutralization assay

Serum samples were collected from re-positive cases, cases in hospitalization and general discharged COVID-19 cases at more than 21 days post illness onset. Microneutralization antibody assays for SARS-CoV-2 were performed in a BSL-3 laboratory according to the standard protocol of a neutralization test. A local SARS-CoV-2 strain isolated from the first COVID-19 patient in Guangdong (GISAID accession ID: EPI_ISL_403934) was used microneutralization assay. All neutralizing antibody assays were run in 96-well microplates. Serum samples were inactivated at 56°C for 30 mins before use, diluted two-fold from 1:4 to 1:1,024, and then incubated at 37°C for 2 hours with equal volumes of 100 half tissue culture infective doses (100 TCID50). Thereafter, the mixture was added into 96-well Vero-E6 cell culture plate. The viral-induced CPE was monitored daily for 7 days. All the diluted samples were tested in duplicate. Cell control, serum control and virus control were included in each plate. Virus back titration was conducted in each test. The antibody titre of the sample was defined as the highest dilution that could inhibit CPE development in 50% of the virus-infected wells.

### High-throughput sequencing

For the multiplex PCR approach, we followed the general method of multiplex PCR as described in (https://artic.network/ncov-2019) (9). Briefly, multiplex PCR was performed with two pooled primer mixtures and cDNA reverse-transcribed with random primers was used as a template. After 35 rounds of amplification, PCR products were collected and quantified, followed by end-repairing and barcoding ligation. Around 50 fmol of final library DNA was loaded onto the MinION sequencing device. The ARTIC bioinformatics pipeline for COVID (https://artic.network/ncov-2019) was used to generate consensus sequences and call single nucleotide changes relative to the reference sequence (MN908947). Assembly of the nanopore raw data was performed using the ARTIC bioinformatic pipeline for COVID-19 with minimap2 (10) and medaka (https://github.com/nanoporetech/medaka) for consensus sequence generation. Sequencing data after mapping to SARS-COV-2 reference genome (MN908947.3) have been deposited in the Genome Sequence Archive (11) in BIG Data Center (12), Beijing Institute of Genomics (BIG), Chinese Academy of Sciences, under project accession numbers CRA002500, publicly accessible at https://bigd.big.ac.cn/gsa.

### Statistical analysis

Statistical analyses were completed using R version 3.5.1 and GraphPad Prism 8.0 (GraphPad Software, Inc., San Diego, CA). Continuous variables that fitted a normal distribution were compared using Student ‘s t-test and analysis of variance (ANOVA) otherwise they were analysed using the Wilcoxon rank test and Kruskal-Wallis H test. Categorical variables were compared using a Chi-square test or Fisher’s Exact Test to assess deviation from the null hypothesis. Spearman’s correlation was to assess the correlation between age and Neutralization antibody titre. A *p*-value <0.05 was determined to be statistically significant.

### Ethics Approval

This study was reviewed and approved by the Medical Ethical Committee of Guangdong Provincial Center for Disease Control and Prevention. Data collection and analysis of cases were determined by the Health Commission of Guangdong province to be part of a continuing public health outbreak investigation during the emergency response and were thus considered exempt from institutional review board approval.

## Results

### Clinical characteristics of re-positive cases

A total of 619 COVID-19 cases were discharged from designated hospitals between 23 January and 19 February, 2020. These cases were continuously isolated in designated hotels and were all screened for SARS-CoV-2 after discharge (see details in Methods). Up to 25 February 2020, 87 cases (14%) retested as positive for SARS-CoV-2 RNA via RT-PCR and returned to the local designated hospital for isolation and medical observation. The demographic characteristics of 87 re-positive cases are as follows: (i) the gender distribution was equal, with 45 males and 42 females; (ii) re-positive detection of viral RNA was observed in all age groups, ranging from 11 months to 68 years, with an average age of 30.4 years, which is significantly younger than that of general COVID-19 patients in Guangdong (average age of 44.8 years, Table 1). Notably, all re-positive cases had only mild (46) or moderate (41) clinical symptoms during initial hospitalization and first retested as SARS-CoV-2 viral RNA positive at a mean of 6.7 days (range 3 to 10 days) post-discharge. Possibly due to their moderate clinical symptoms, re-positive cases had shorter hospital stays (mean, 14.8 days) than general COVID-19 cases (mean, 20.1 days) (Table 1). After discharge, 77 of 87 re-positive cases were asymptomatic, and 10 cases had a symptom of unproductive cough, mainly at night. Forty-four cases received computerized tomography (CT) examination, and no abnormalities were reported.

**Table 1.** Demographic and clinical data of re-positive and normal COVID-19 cases

|  | Normal cases | Re-positive cases | *P* value |
| --- | --- | --- | --- |
| Demographics |  |  |  |
| Age (average, range) | 44.8 (0.8-90) | 30.4 (0.9-68) | <0.001 |
| Gender |  |  |  |
| Male | 218/425(48.7%) | 45/87(51.7%) | >0.05 |
| Female | 207/425(51.3%) | 42/87(48.3%) |  |
| Clinical classification |  |  |  |
| Mild | 28/256(10.9%) | 46/87(52.9%) | <0.001 |
| Moderate | 167/256(65.2%) | 41/87 (47.1%) |  |
| Severe | 61/256(23.8%) | 0/87(0) |  |
| Clinical course (n=69) |  |  |  |
| Onset-hospitalization (days) | 6.7 (0-37) | 3.2 (0-13) | <0.001 |
| Hospital stay | 20.1(4-32) | 14.8 (6-28) | <0.01 |
| Discharge-sampling | N/A | 6.7 (3-10) | N/A |
| Onset-discharge | N/A | 18.8 (9-37) | N/A |
| Onset-sampling | N/A | 33.2 (19-47) | N/A |

Note: N/A indicates unavailable data. Clinical data were from 69 re-positive cases since these data were incomplete among 87 re-positive cases.

### Neutralizing antibody in re-positive cases

An impaired immune response has been associated with fatal COVID-19 infections that exhibit prolonged persistence of viral RNA (13). One possible explanation for the re-positive detection of SARS-CoV-2 RNA is that some COVID-19 patients may have insufficient immune responses and neutralization antibodies (NAbs) to clear infection completely. To investigate the immunological and virological characteristics of re-positive COVID-19 cases, 70 of 87 re-positive cases were resampled by Guangdong Provincial Center for Disease Control and Prevention (GDCDC) between 22 February and 1 March, 2020 including 59 serums and 137 swab samples (Figure 1A). Serum samples were collected at a median of 35 days post illness onset (ranging from 23 to 47 days). As a comparation, 150 serum samples from general discharged cases (n=38) and cases in hospitalization (n=112) were collected with a median duration from illness onset to serum sampling of 30 days (ranging from 22 to 47 days) and 32 days (ranging from 22 to 45 days), respectively. The titre of viral-specific NAbs was estimated by microneutralization assay, which is regarded as a gold standard for determining protective antibodies. As shown in Figure 1B, 58 of 59 (98.3%) re-positive cases developed NAbs with a titre >4, ranging from 4 to >1024. Our results demonstrated competent immune activation in re-positive cases, which exhibit a distribution of NAbs titres similar to that of general discharged cases and COVID-19 cases in hospitalization (Kruskal-Wallis H test, *p*=0.12, Figure 1B).

**Figure 1.**
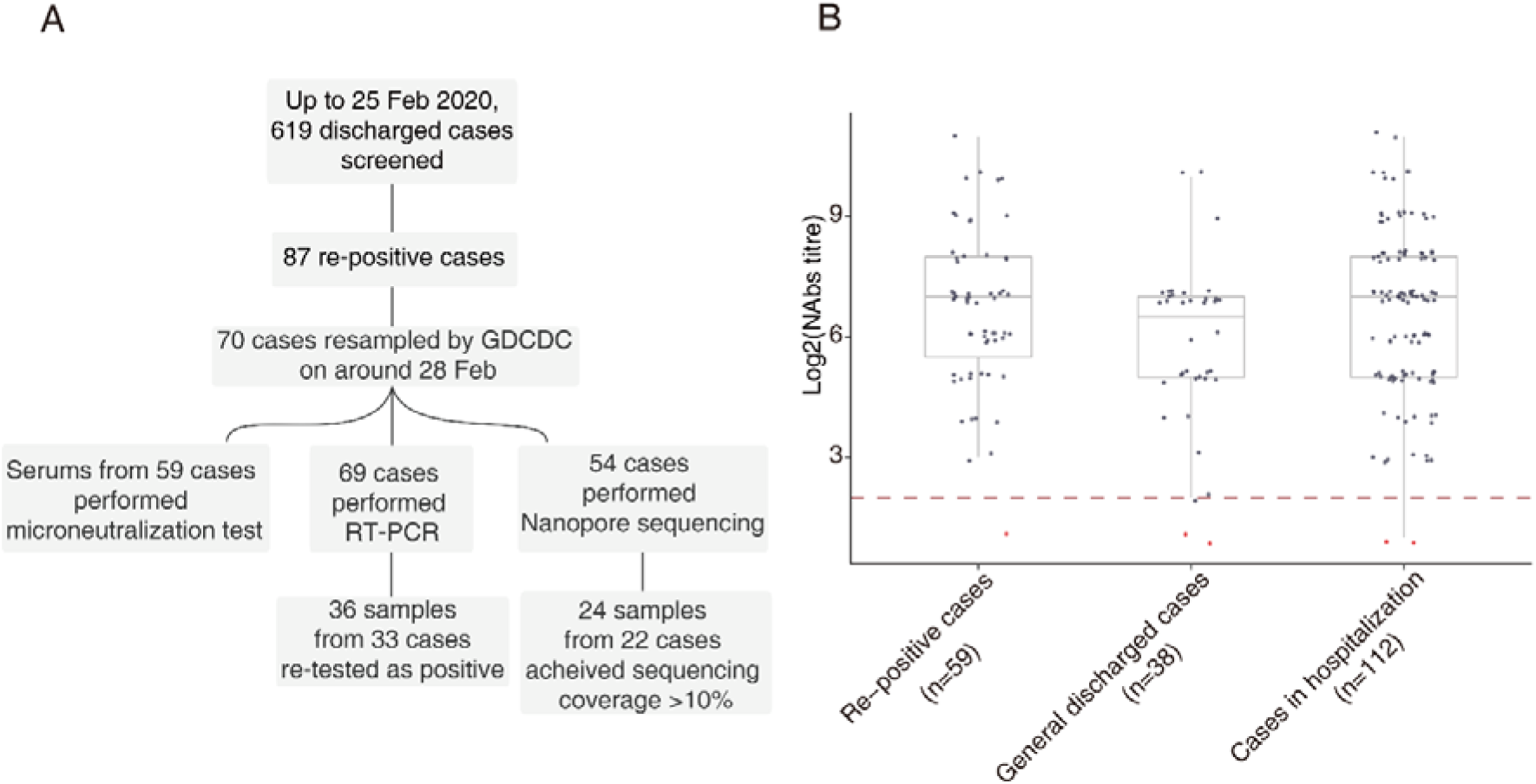
(A) Summary of sampling scheme in this study; (B) Comparison of Nab titres among infections that were re-positive, general discharged COVID-19 cases, and cases in hospitalization. General discharged cases refer to COVID-19 recovered cases detected as SARS-CoV-2 negative in 14 days after discharge. To plot these results into one figure, we recorded 2 for antibody titre less than 4 (highlighted in red), and 2048 for titre larger than 1024. The start point of serum dilution (titre of 4) was highlighted with red dash line.

### Viral RNA detection and viral isolation in re-positive cases

A total of 137 swabs, including 51 nasopharyngeal swabs, 18 throat swabs and 68 anal swabs, were tested using three different RT-PCR kits, in an attempt to reduce the chance of false negatives caused by difference in sensitivity and primer specificity. Fifty re-positive cases had paired nasopharyngeal swabs and anal swabs, and 18 cases had paired throat swabs and anal swabs for viral RNA detection. Thirty-six swabs from 33 cases were detected as positive by at least one RT-PCR kit (Table S1). RT-PCR positive rates were not statistically different for different sample types in 68 paired samples (anal swabs vs nasopharyngeal swabs, and anal swabs vs throat swabs; Chi-square test, *p*=0.648; Fisher exact test, *p*=0.443). In this cross-sectional analysis, 32 of 33 cases had clear information on time intervals of illness onset, discharge and sample testing (Figure 2). We find that the duration from discharge to the time tested as re-positive can range from 6 to 28 days (Figure 2). Importantly, there was no correlation between neutralization antibody titre and the length of time between discharge and the date the case tested as re-positive. For example, for Case 21, SARS-CoV-2 RNA was re-detected at 28 days after discharge and 46 days post symptom onset, yet NAb titre for this case was as high as 1024 (Figure 2). The 36 RT-PCR positive samples including 14 nasopharyngeal swabs, 3 throat swabs and 19 anal swabs. These RT-PCR positive samples were inoculated into Vero-E6 cell line but no live viruses could be cultured following two rounds of cell passage.

**Figure 2.**
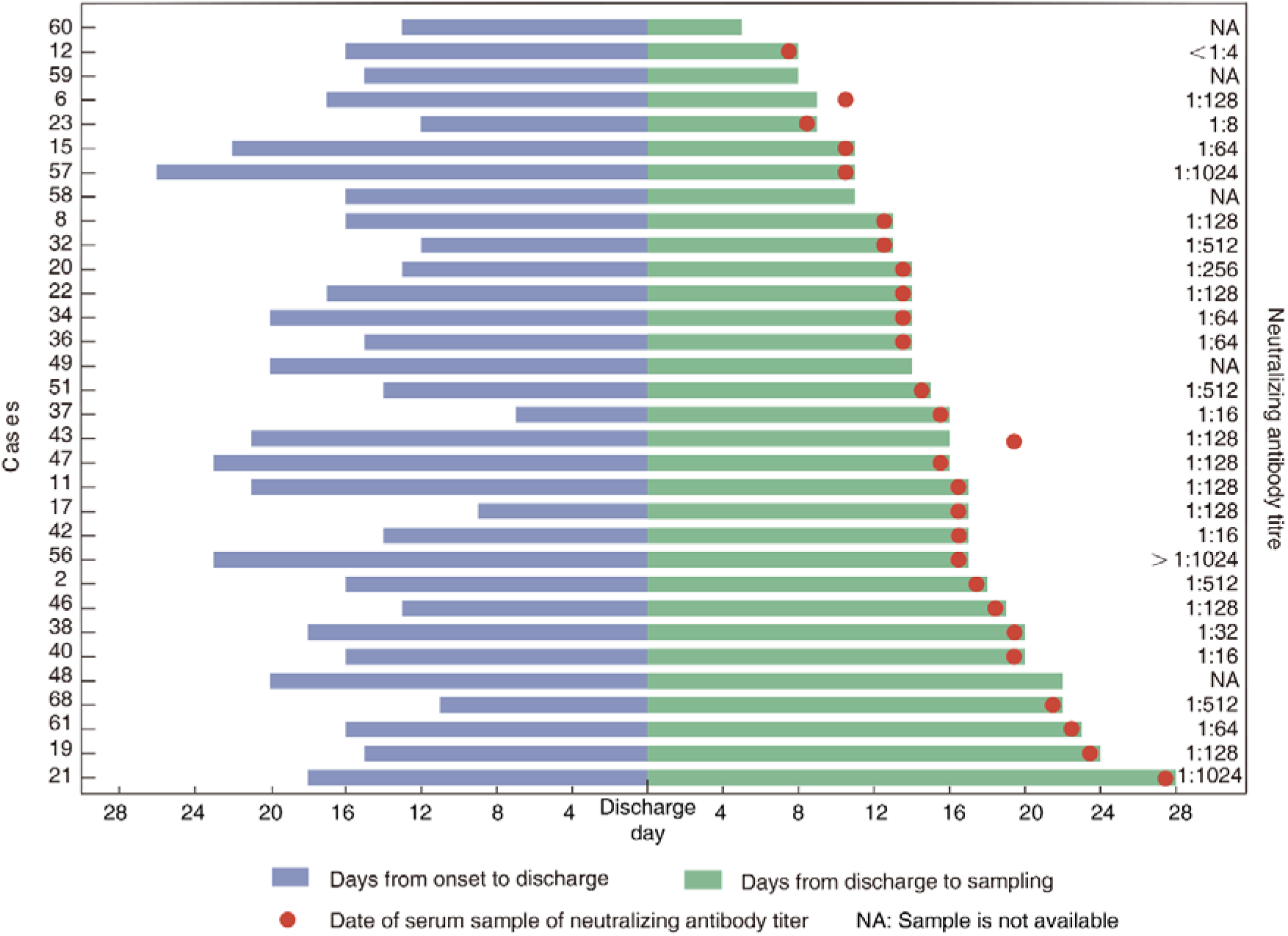
Timeline of 32 COVID-19 re-positive cases sampled and tested between 28 February–1 March.

### Virus whole genome sequencing in re-positive cases

A previously study proved virus isolation success was also depended on viral load, and samples containing <10^6^ copies/mL (or copies per sample) never yielded an isolate (3). For acute infection cases, we found that high quality SARS-CoV-2 genomes could be obtained by using a multiplex PCR method even for samples with a low viral load (8). Therefore, the successive detection of a complete or nearly complete viral genome may provide clues on the status of viral replication. None of full-length SARS-CoV-2 genome could be obtained by sequencing 94 samples from 54 patients and the sequencing coverage ranged from 0.00–75.48% (Figure 3A, Table S1).

**Figure 3.**
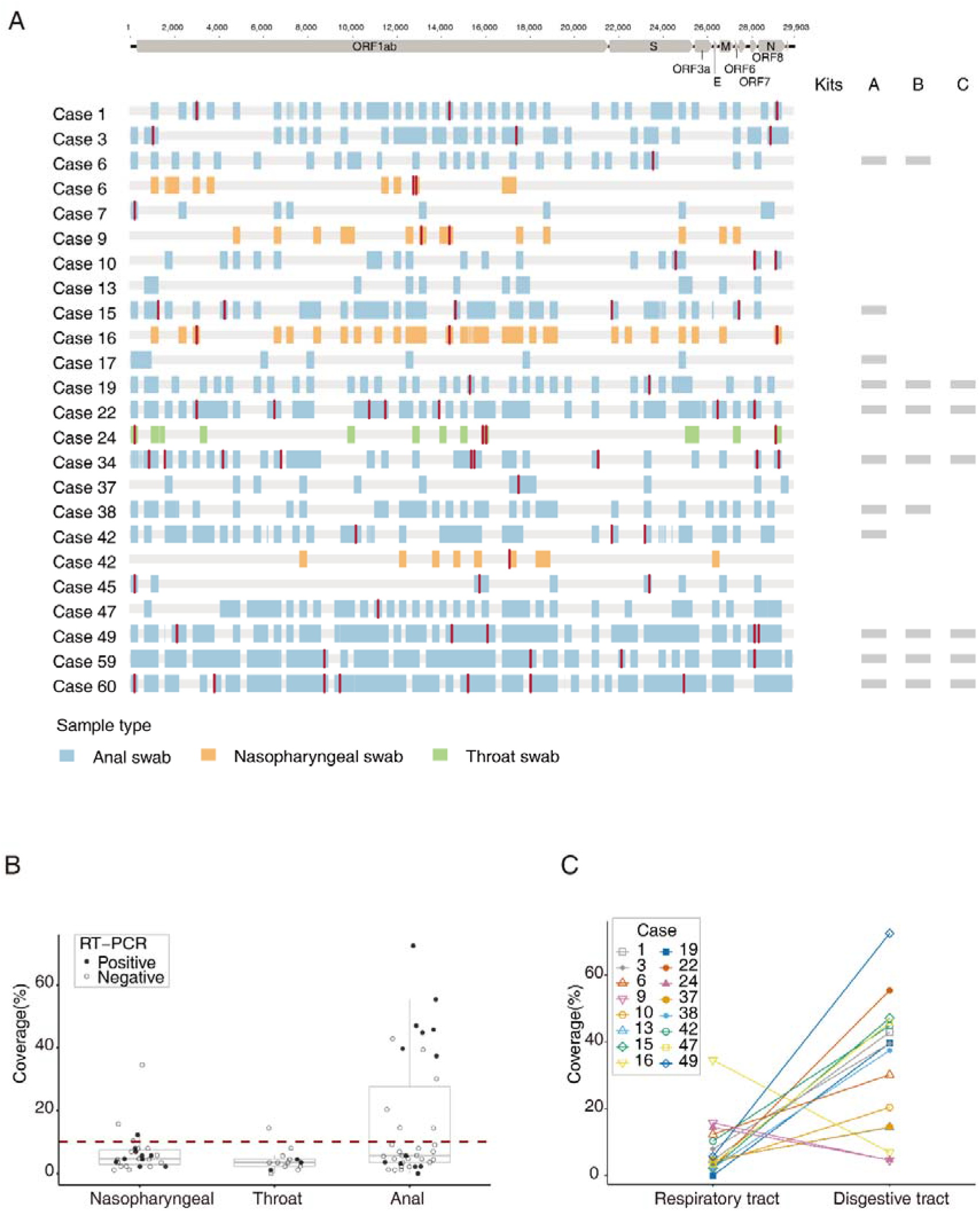
(A) Virus genome sequences (for those samples with genome coverage >10%) obtained from re-positive cases. Single nucleotide polymorphisms (with respect to the reference genome MN908947.3) are coloured in red. Genome sequence fragments are coloured blue, orange to indicate whether they were obtained from anal, nasopharyngeal and throat swabs, respectively. The corresponding RT-PCR results from three different RT-PCR kits were shown on the right side and positive results were marked with rectangles. (B) Coverage of the consensus sequence among nasopharyngeal swabs, throat swabs and anal swabs. A solid circle refers a RT-PCR positive sample, and a hollow circle refers a RT-PCR negative sample. The red dash line refers to the sequencing coverage of 10%; (C) Coverage of consensus sequence measured from respiratory tract (nasopharyngeal swabs/throat swabs) and matched digestive tract (anal swabs).

Intriguingly, a discrepancy was observed between the results of RT-PCR and multiplex PCR sequencing (Figure 3A & B). For instance, 21 of 33 samples that were RT-PCR positive did not perform well in sequencing and produced sequences that covered < 10% of the virus genome (Figure 3B). Conversely, 12 samples detected as negative by three RT-PCR kits gave rise to virus sequences that spanned >10% of the virus genome (Figure 3A & 3B). This discrepancy could be expected when the viral genome was not intact and the primers in RT-PCR and multiplex PCR targeting the different fragments of viral genome.

Sequencing results of matched samples from individual cases (anal swabs vs nasopharyngeal swabs, and anal swabs vs throat swabs) showed that the genome coverage of sequences from digestive samples were significantly higher than that of the matched respiratory swabs (Figure 3C). For case 6 and case 42 having sequencing results of matched samples, we observed some single nucleotide variants (SNVs) between viral genome sequences achieved from respiratory samples and that from digestive samples (Figure 3A).

## Discussion

Tens of thousands of people have recovered from COVID-19 infection, and there have been preliminary reports of people testing re-positive for SARS-CoV-2 viral RNA after recovery [7,8]. Here, we use data from the Guangdong COVID-19 surveillance system to analyse the characteristics of re-positive cases in Guangdong between 23 January and 26 February, and we explore questions such as the percentage, immune status and infectivity of re-positive cases.

The first question we addressed is the re-positive rate in COVID-19 discharged cases. Here, the discharge criteria are according to the Diagnosis and Treatment Scheme of SARS-CoV-2 (see Methods for detail). By screening 619 discharged cases, up to 25 February, the percentage of SARS-CoV-2 re-positive cases is around 14%. According to the scheme, all discharged cases are continuously isolated in designated hotels with strict interventions on diseases transmission. Thus, the identification of re-positive SARS-CoV-2 during the isolation phase exclude the possibility that re-positive cases is caused by the secondary viral infection. Our results also highlight a significant feature of re-positive cases. All re-re-positive cases in our study developed only mild or moderate symptoms in the initial diagnosis, with the median age significantly lower than that of the general COVID-19 cases (Table 1). The relatively mild symptoms may explain why the median time from onset to discharge in re-positive cases (median 19.3 days) is slightly lower than that of the other discharged COVID-19 cases (median 24 days). It is unlikely that cases tested as re-positive because they were discharged too early since all re-positive cases tested negative for both nasopharyngeal and anal swabs in two successive tests before discharge. The time from symptom onset to discharge (the time when COVID-19 cases have twice tested negative by PCR) for re-positive cases (median 19.3, Table 1) is consistent with the time that detectable SARS-CoV-2 RNA is reported on other studies to persist (median 20) in respiratory sites (13, 14). These data indicate that the course in re-positive cases is similar to other COVID-19 cases. The re-positive of SARS-CoV-2 RNA is not random and mainly observed in young cases without severe clinical symptoms.

Prolonged detection of virus RNA presents a challenge to targeted public health interventions. Therefore, it is important to know if re-positive cases are infectious. One previously proposed reason for prolonged detection of viral RNA in deceased patients is impaired neutralizing ability (13). Our microneutralization result shows that 58 of 59 (98%) re-positive cases generated specific NAbs to SARS-CoV-2, and their titre distribution is similar to the normal recovered cases and hospitalized COVID-19 cases (Figure 1B). To investigate virus infectivity, we attempted live virus isolation on different clinical samples from re-positive cases. All samples from re-positive cases are diagnosed with higher CT values (Table S1) than our previous finding in acute infection cases. Due to the limited sensitivity of culture method (3), infectivity may not accurately illustrated by the culture method although no viral isolates were obtained from RT-PCR positive samples. Thus, we also perform the multiplex PCR combined with high-throughput sequencing on these samples. The discrepancy we observed among different RT-PCR kit results, as well as between RT-PCR and multiplex PCR sequencing results, suggests that the virus genomes detected in re-positive cases could be highly degraded. We only recovered one virus sequence with genome coverage >20% (34.5%) in 23 RT-PCR positive respiratory samples (Figure 3A). These results suggest a low residual risk of infectivity of re-positive cases, especially from the respiratory transmission route.

Several limitations of our study should also be noted. First, we did not obtain successively collected samples, resulting in the existence of bias toward the summarized duration from the discharge to firstly re-positive result for viral RNA as well as the time of the re-positive RNA to negative. Secondly, we did not obtain the corresponding samples during the acute infection for these re-positive cases. Therefore, some virologic questions remain, including whether there any genetic differences for SARS-CoV-2 viruses sampled in an acute infection phase and a re-positive phase. The significance of SNVs identified in different samples of re-positive cases is limited by the sample size (Figure 3A) and should be further clarified in following studies.

Appropriate design intervention strategies on COVID-19 has been largely relied on how well we understand the characteristics of the SARS-CoV-2 infection. Re-positive of viral RNA in some discharged cases could raise a challenge for disease interventions which means a prolonged isolation phase and a more requirement on hospital isolation facilities. Our study result shows a comparable Nab response in re-positive cases comparing to other COVID-19 cases. More importantly, none of infective strains could be successfully isolated and no intact viral genome could be sequenced from all re-positive cases samples highlighting a lower risk for disease transmission from such cases. The additional educations related on SARS-CoV-2 re-positive should be performed to calm down the public panic and allocate of limited medical resources.

## Data Availability

The data used to support the findings of this study are available from the corresponding author upon request.

## Acknowledgments

We thank the laboratory and administrative personnel at Guangdong Provincial Center for Disease Control for their contribution to the follow-up investigation. We also acknowledge the contributions of other clinical, public health and technical staff from COVID-19 designated hospitals and city-level center for disease control and prevention. This work was supported by grants from Guangdong Provincial Novel Coronavirus Scientific and Technological Project (2020111107001), Science and Technology Planning Project of Guangdong(2018B020207006), National Science and Technology Project(2020YFC0846800).

